# Determinants of catastrophic health expenditure attributable to non-communicable diseases and impoverishment in Pakistan

**DOI:** 10.1101/2022.11.28.22282844

**Authors:** Lubna Naz, Shyamkumar Sriram, Filzah Sardar

## Abstract

**Background:** Pakistan has a disproportionately high burden of non-communicable diseases (NCDs), leading to an increase in healthcare utilization and associated out-of-pocket health expenditure, adversely affecting the well-being of the household. This study aims to identify the determinants of catastrophic health expenditure (CHE) on NCDs and quantify the impoverishment effects of OOP expenditure attributable to NCDs.

**Methods:** The study used Household Integrated Economic Survey - 2018/2019 and the National Health Accounts Data 2017. The welfare impact of out-of-pocket health spending associated with NCDs was assessed using specific measures: a) incidence and intensity of catastrophic health expenditure and b) the impoverishing effect. A generalized linear model with a logit link function was used to study the determinants of CHE at different thresholds.

**Results:** The poverty headcount was 20.5% without accounting for OOP expenditure for NCDs; with adjustment, it increased to 27%, causing 13 million (from 42.4 million to 55.6) more people to fall into poverty. Households experiencing CHE fell from 60% to 3.5% as the threshold increased from 10% to 40%, implying fewer households encounter CHE at higher thresholds. Larger families, male-headed, families with children and older members, having more members with NCDs, and using private healthcare were more likely to incur CHE.

**Conclusions:** CHE has a high propensity to push households into poverty. Pakistan’s National Health Vision 2016-2025 recognizing the provision of Universal Health Coverage and poverty alleviation as the top health and social priorities needs to be implemented to achieve Sustainable Development Goal targets of UHC and financial risk protection.

**Key messages:**

- Household’s out-of-pocket spending associated with non-communicable disease was USD19 per month.
- Female headed families had a lower likelihood of incurring catastrophic health expenditure on NCDs than the male.
- Only a fewer households encountered catastrophic health expenditure at higher thresholds.
- Rural households had a higher impoverishing effect of out-of-pocket expenditure associated with NCDs compared to the urban.

## Introduction

Traditionally, non-communicable diseases (NCDs) were considered to affect high-income countries (Tchewonpi et al. 2013). Even a few decades ago, non-communicable diseases (NCDs) were considered an evolving epidemic in developing countries; however in the current scenario, the incidence and financial burden associated with NCDs in developing countries are increasing at an alarming rate (Ndubuisi 2021).

Globally, non-communicable diseases contribute to around 35 million deaths, accounting for 60% of all deaths worldwide. About 80% of these deaths occur in low- and middle-income countries (Ndubuisi 2021). An increase in the number of risk factors, such as a sedentary lifestyle, growth in the consumption of unhealthy diet, tobacco, and uncontrolled alcohol use in developing countries, has led to the NCD epidemic in these countries (Ndubuisi 2021). Among the NCDs, the three major diseases, cardiovascular diseases, diabetes, and cancer, constitute a sizeable fraction of the mortality in the Asia-Pacific Region (Low, Lee, and Samy 2015).

In Pakistan, non-communicable disease and injuries constitute over 77% of age-standardized deaths (Lozano et al. 2012). Specifically, the mortality due to cardiovascular diseases and diabetes in Pakistan was about 455 per 100,000 among the male population and 388 per 100,000 among females (Low et al. 2015). Similarly Pakistan had higher mortality due to cancer, with 95 deaths per 100,000 males and 94 deaths per 100,000 females (Low et al. 2015). By 2025, it is estimated that around 3.87 million premature deaths will be caused by NCDs in Pakistan (Rafique et al. 2018). The nature and severity of NCDs-related illnesses have severe health outcomes and economic consequences for the individual and household. Over the past decade, there has been increasing attention on the policies to reduce the burden of NCDs globally and to provide financial protection to the population for the utilization of healthcare services. The third high-level meeting on the control of NCDs was conducted by the United Nations in September 2018 to track the progress towards the commitments made for the control of NCDs and to provide Universal Health Coverage (UHC) to the population (Murphy et al. 2020).

Financial protection is considered to be one of the goals under UHC(Tchewonpi et al. 2013). The chronic nature of NCDs has led to increased healthcare utilization for treating chronic diseases and their associated life-long complications. The protracted effect of NCDs leads to the need for long-term care by incurring catastrophic health expenditure which are likely to push households into poverty. The United Nations has pledged to reduce premature mortality due to NCDs and the associated higher costs of treating NCDs, which create an undue burden on people already suffering from chronic illnesses (Murphy et al. 2020).

The World Health Report 2010 mentions the three dimensions of UHC, namely population covered, cost, and service (WHO 2010). Achieving UHC is also one of the critical targets under the Sustainable Development Goals (SDGs). UHC cannot be achieved without providing financial protection to the population by eliminating catastrophic health expenditure and the associated impoverishment due to OOP health expenditure. The SDGs aim to reduce premature mortality from NCDs through prevention and treatment, promote mental health and well-being, and achieve Universal Health Coverage (UHC), including financial protection. Specifically Target 3.8 of the SDG aims to provide “Achieve universal health coverage, including financial risk protection” (Asia, 2017). In addition, SDG 1 and 5 aim to end extreme poverty and inequality (Thomson, Mosca, and Evetovits 2020).

With the rapid epidemiological transition in many developing countries, including Pakistan, the incidence and prevalence of NCDs are rising compared to the burden of traditional infectious diseases that were earlier common in the region (Blum, Majid, and Hotez 2018). This high burden of NCDs and the associated healthcare utilization and costs make households spend a higher proportion of their disposable income on treating these illnesses. There were many studies done globally that showed that the costs of treatment and care for NCDs deprive households of the essential resources that could otherwise be used for other vital needs like education or food for the family (Gertler and Gruber 2002; Mwai and Muriithi 2016).

Out-of-pocket payment for healthcare tends to push households into poverty or deepen the poverty of already poor households (Xu et al. 2003). Evidence also showed that poor people in developing countries refrain from obtaining treatment for NCDs and have poor adherence to care because of the fear of incurring OOP costs (Huffman et al. 2011; Murphy et al. 2013, 2016, 2018; Tchewonpi et al. 2013). Costs associated with NCDs include not only the costs of treatment of NCDs but also the costs of loss of income and working time associated with lost productivity due to both premature mortality and the long duration of disability due to chronic illnesses (Tchewonpi et al. 2013). Many developing countries only have limited health insurance coverage for NCDs and their complications. Thus, the significant financial burden of paying for care falls on households, exposing them to financial distress (Tchewonpi et al. 2013).

Pakistan is currently making a policy change and taking initiatives to provide UHC to its population. The latest initiative is the Government of Pakistan announcing a health card for the entire population of the most populated province of Punjab after the initiative was piloted in the Khyber Pakhtunkhwa province (Farooq, Arshad, and Usman 2022). The health card covers the cost of hospitalization for several chronic diseases (Farooq et al. 2022). Although Pakistan is going through several rapid policy changes and reforms to provide UHC and reduce OOP health expenditure, there is little evidence that OOP expenditure are catastrophic to the population due to NCDs and their role in pushing households into poverty.

Prior studies have either explored the determinants of OOP health expenditure (Malik 2015) or the determinants of CHE and impoverishment in Pakistan (S.Bashira, S.Kishwar, and Salman 2021; Shujaat Farooq and Masud 2021) or have analyzed the trends in OOP health expenditure over time (Khalid and Sattar 2016), no study so far has focused specifically on the OOP-related financial burden due to NCDs. Therefore, identifying the degree of impoverishment and extent of CHE due to NCDs will help policymakers design adequate health insurance programs and other mechanisms to address and mitigate the financial burden. Thus, this study aims to contribute to the literature by identifying determinants of incurring CHE associated with NCDs among households in Pakistan and quantifying impoverishment effects of OOP spending on NCDs.

## Methods and Material

### Data sources

This study extracted the data of 14,413 households, comprising at least one member inflicted with NCDs in the past three months prior to the survey, from the Household Integrated Economic Survey (HIES) -2018/2019. The survey was implemented by the Pakistan Bureau of Statistics (PBS) from August 2018 to June 2019. A two-stage stratified random sampling design has been adopted. PBS has collected micro-data through Pakistan Social and Living Standards Measurement Survey (district level) and Household Integrated Economic Survey (national and provincial level) on alternating years since 2004. So far, seven rounds have been collected, ranging from 2004/05 to 2018-19.

HIES 2018-19 contains information on 24,809 households from all over Pakistan, including 15,936 rural and 8,873 urban households. The current study used information on socio-demographic characteristics of households, education, employment, self-reported health, healthcare providers, types of disease and treatment, food and non-food expenditure, and income. HIES 2018-19 is available in the public domain at https://www.pbs.gov.pk/content/pslm-hies-2018-19-microdata. Furthermore, HIES was complemented with the National Health Accounts Data obtained from PBS. The latter provided a detailed account of OOP health expenditure on NCDs.

### Conceptual Framework

This study followed the analytical frameworks proposed by Li et al., (2012) and Kumara and Samaratunge (2016), which postulate that families’ capacity to incur CHE (OOP health spending on NCDs as a fraction of total household expenditure exceeding a certain limit) depends on a range of factors which can be broadly categorized into demand-side and supply-side determinants. The demand-side factors include socio-demographic characteristics of the household, head’s age, marital status and education, family’s income, members inflicted with NCDs, and age composition of the family, see Figure 1. The supply-side factors generally include the access and availability of healthcare, healthcare financing, and access to social safety nets. In Pakistan, healthcare insurance is generally available to people in the formal sector. The healthcare financing program (Sehat Sahulat program, SSP) launched in 2015 is primarily restricted to only two provinces, Khyber Pakhtunkhwa, and Punjab. It was due to disagreement between federal and provinces over the execution and monitoring of the program, as aftermath the 18th amendment in the constitution, provinces have autonomy in the healthcare system management (Hasan et al. 2022).

**Figure 1.**
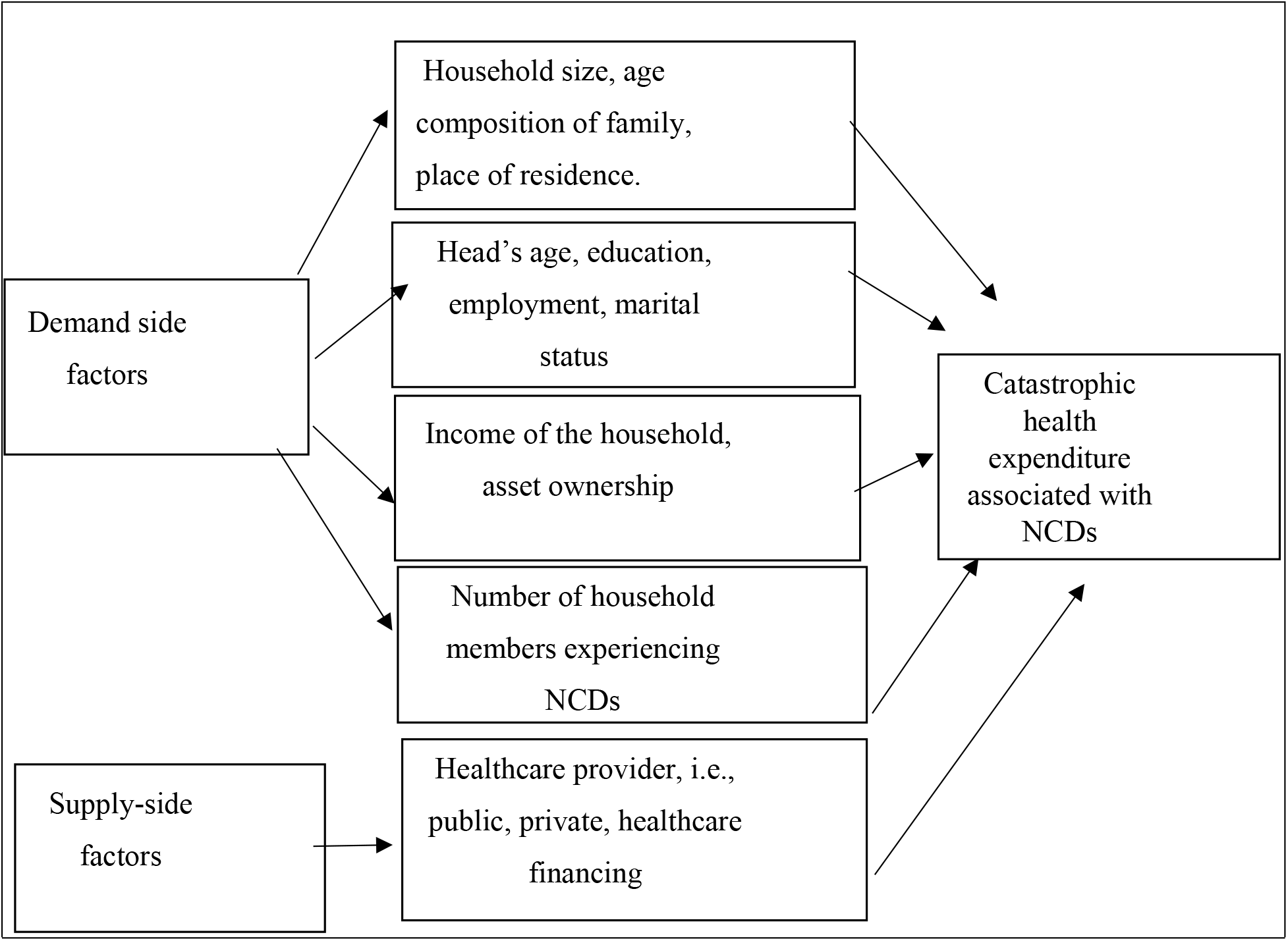
Diagrammatic framework of the determinants of CHE attributable to NCDs

### Empirical methodology

To assess the welfare impact of OOP health spending associated with NCDs, we used the following measures, a) incidence and intensity of CHE attributable to NCD, and b) the impoverishing effect (i.e., poverty headcount, poverty gap, and poverty squared gap) of CHE associated with NCDs. Finally, we estimated the model, as set up under the analytical framework, of the determinants of CHE attributable to NCDs. The OOP expenditure on NCDs was calculated by aggregating medical and non-medical expenses. Data on doctor’s fees, medicine, diagnostic tests, surgery cost, medical supplies, and other durables such as oxygenated blood made up medical expenses. Information on transport costs, food, admission fees, and accompanying personal costs constituted the non-medical expenses. The total household expenditure comprised food and nonfood expenditure, excluding food eaten away from home, food eaten at weddings or in social gatherings, license fees, taxes, and others.

Finally, the share of OOP spending associated with NCD treatment was computed as a fraction of the total household expenditure exceeding the 10% threshold, whereas varying thresholds of 5%, 15%, and 20% were used for sensitivity analysis. Generally, the lower thresholds (i.e., 5% or 10%) imply a fraction of total household consumption or income. In contrast, a noticeably higher one (i.e., 40%) is calculated as a ratio of OOP expenditure on health to nonfood expenditure (O’Donnell et al. 2008). The following formula was used for calculating CHE on NCDs:

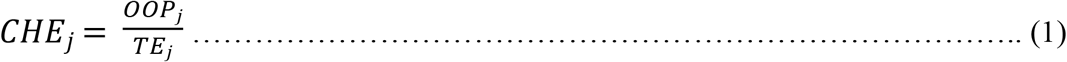

Where OO*Pj* represents *j* household’s out-of-pocket expenditure on NCDs and *TEj* represents the total expenditure of households. Consider *CHEj* as the proportion of household *j*’s healthcare spending and *T* as the threshold (10%) after which household *j* underwent catastrophic spending if CHE*j* > *T*. We calculated the incidence of CHE on non-communicable diseases using a 10%, 15%, 20%, and 40% thresholds of non-food expenditure, as follows,

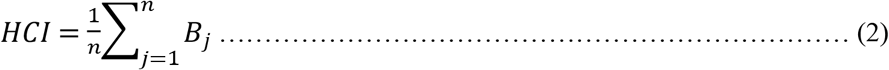

Where the sample size is represented by n and *B*_*j*_ is a measure equal to 1 if *CHE*_*j*_ > T and 0 otherwise. Equation 2 implies the percentage of households whose OOP healthcare expenditure on NCDs surpass the 10%, 15%, 20%, and 40% thresholds in the reference period. However, the headcount index (HCI) does not reveal the intensity of CHE, i.e., how far the households went over the threshold. Therefore, the study calculated the mean overshoot as follows.

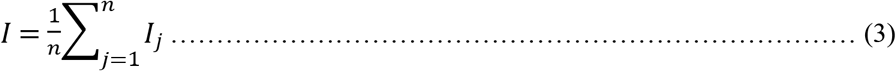

Where *I*_*j*_ is the quantity with which household *j*’s share of OOP health spending crossed the specified threshold, and can be expressed as:

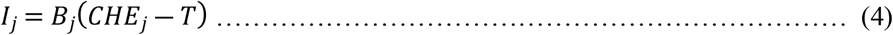

The following equation represents the average positive overshoot.

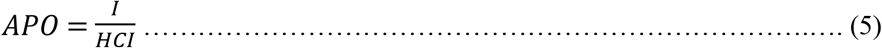

As a result, the catastrophic intensity was equal to the proportion of total expenditure that were catastrophic times the average positive overshoot; I = HCI × APO (Verma, R., Kumar, and Dash 2021).

To calculate the impoverishing effect, firstly, we compared per-adult equivalent household expenditure without accounting for NCDs with the poverty line; the latter was estimated by the World Bank using cost of basic needs approach-CBN in Pakistan, which counts a household poor when its per-adult equivalent consumption falls beneath the poverty line PKR 3,030,PKR 3741 at urban prices and PKR 3,769 at rural prices, in 2018-19 (World Bank 2021). The prepaid poverty headcount ratio was obtained as follows,

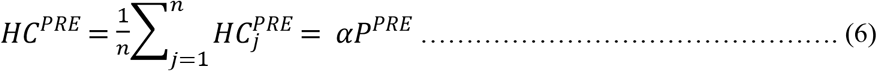

In Equation 6, n represents the adult-equivalence or size of the households and *αP*^*PRE*^ is the proportion of the population that was poor, and *HC*^*PRE*^ =1 if per adult-equivalent monthly spending (*TE*_*j*_) < Poverty line ‘pl’ and 0 otherwise. Households’ OOP payment OO*P*_*j*_ on NCDs was subtracted from prepaid expenditure (*TE*_*j*_ − OO*P*_*j*_) to compute poverty indices for postpaid cost. The impact of OOP expenditure on poverty was then calculated by subtracting prepaid indices from postpaid indices (Verma, R. et al. 2021; Wagstaff, A., & Doorslaer 2003; Wagstaff, A., O’Donnell, O., Van Doorslaer, E., & Lindelow 2007), as given below:

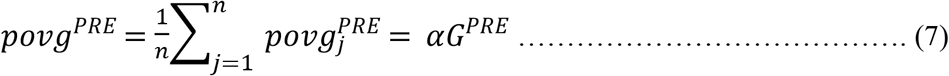

Nevertheless, the difference in poverty incidence revealed the impoverishing effect of OOP expenditure on NCDs; there was still a lack of information on the intensity of poverty. It means the computed measure did not account for how much an impoverished household dropped deeper into poverty. Therefore, we computed the normalized poverty gap to identify the amount by which out-of-pocket spending on NCDs drove a family into poverty.

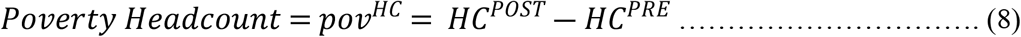

Here, 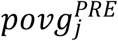 is the prepaid poverty gap, which corresponds to (pl − *TE*_*j*_) if the condition (*TE*_*j*_) < Poverty line pl is satisfied, and 0 if it is not.

Finally, we used the Generalized Linear Model, with Logit as a link function from binomial family to estimate CHE at 5%, 10%, 15%, and 20% thresholds of total expenditure. Using a cue from the literature, a set of explanatory variables, such as region, educational status of head, household size, members with NCDs, marital status of head, head’s employment, age composition of family, gender, and income quintiles were used.

## Results

### Incidence of non-communicable diseases by income quintiles and region

In Pakistan, the prevalence of NCDs shows an inverse relationship with income quintiles. The percentage of families with at least one member experiencing NCDs decreases as one moves from a lower quintile to a higher quintile, see Table 1. The highest percentage of households with NCDs was found in the first three income quintiles (approximately 70%). Across regions, the rural had a higher incidence of NCDs than the urban. Amongst NCDs, the prevalence rate of heart disease was the highest (9.73%), followed by flu/fever (9.55%), muscular pain (such as knee, arm, backbone) (7.21%), and diabetes (7.09%). The second most prevalent NCDs were liver, kidney, hepatitis, and gynecological problems.

**Table 1.**
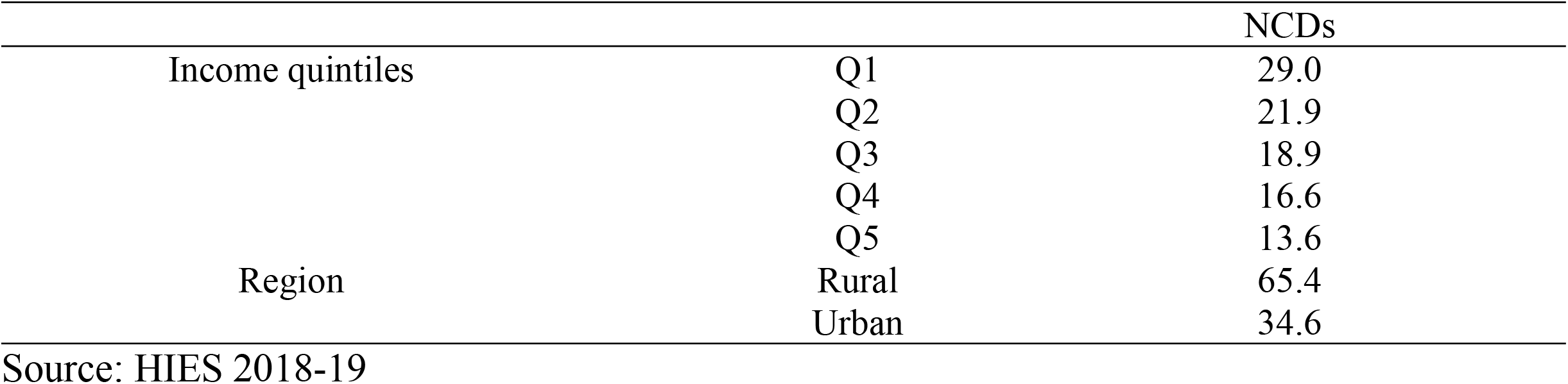
Prevalence of non-communicable diseases by income quintiles and region

Unlike the results of the prevalence rate of NCDs by income quintiles, the out-of-pocket health expenditure by households in the upper quintile was higher than those in the lower quintile, reflecting that wealthier households spent more on healthcare services than poorer households in 2018-19, see Figure 2. It may be due to the frequent healthcare seeking behavior of richer households from private hospitals, which are generally expensive. Poor are not involved in frequent medical consultations, mainly outpatient, due to their limited financial resources and the unavailability of healthcare financing in Pakistan. Moreover, the OOP spending on NCDs was higher in each quintile than the OOP expenditure on healthcare. It reflects a disproportionally higher financial burden of NCDs in Pakistan across all income groups.

**Figure 1.**
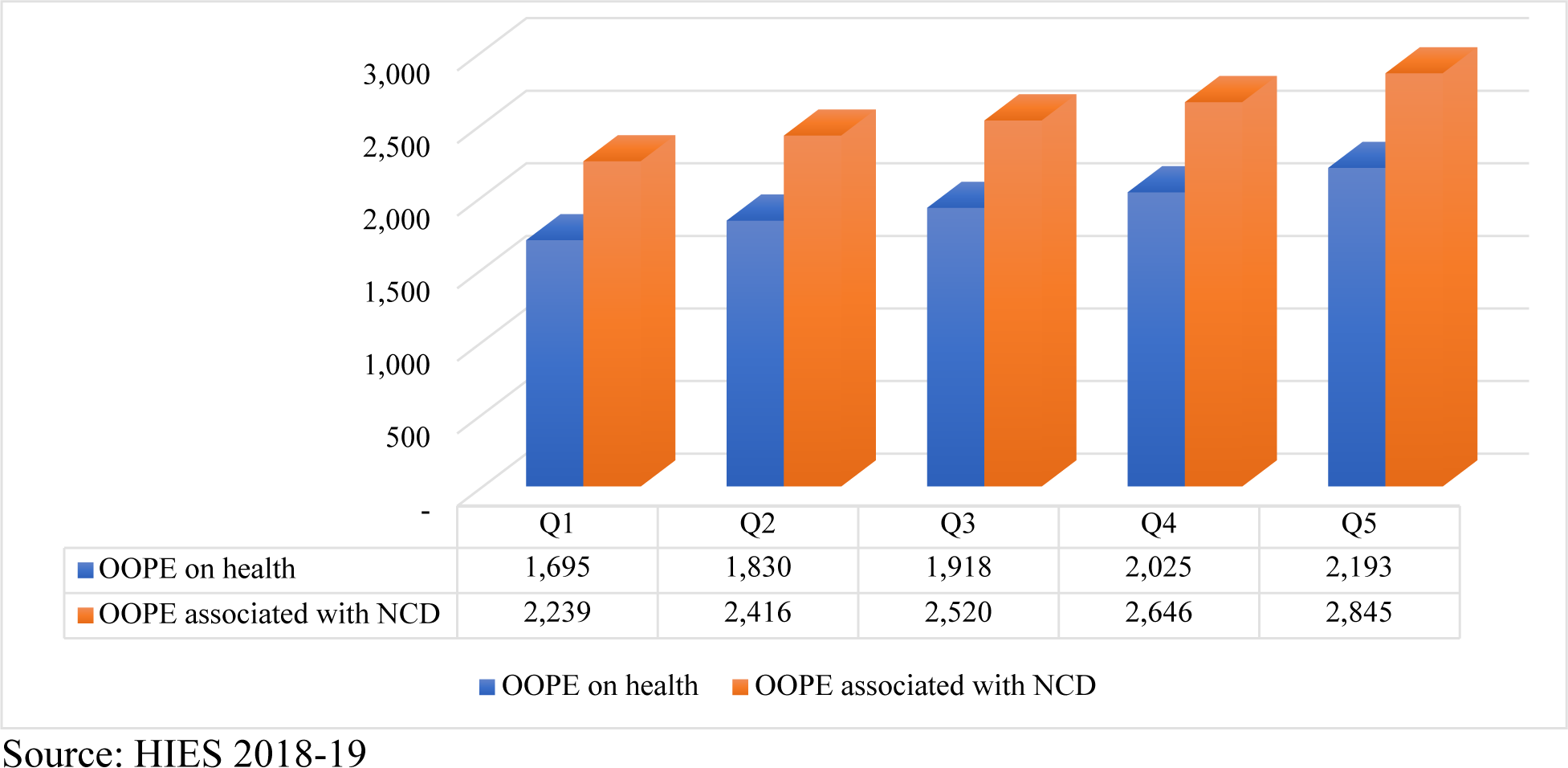
Out-of-pocket expenditure on healthcare and NCDs by income quintiles

**Figure 2.**
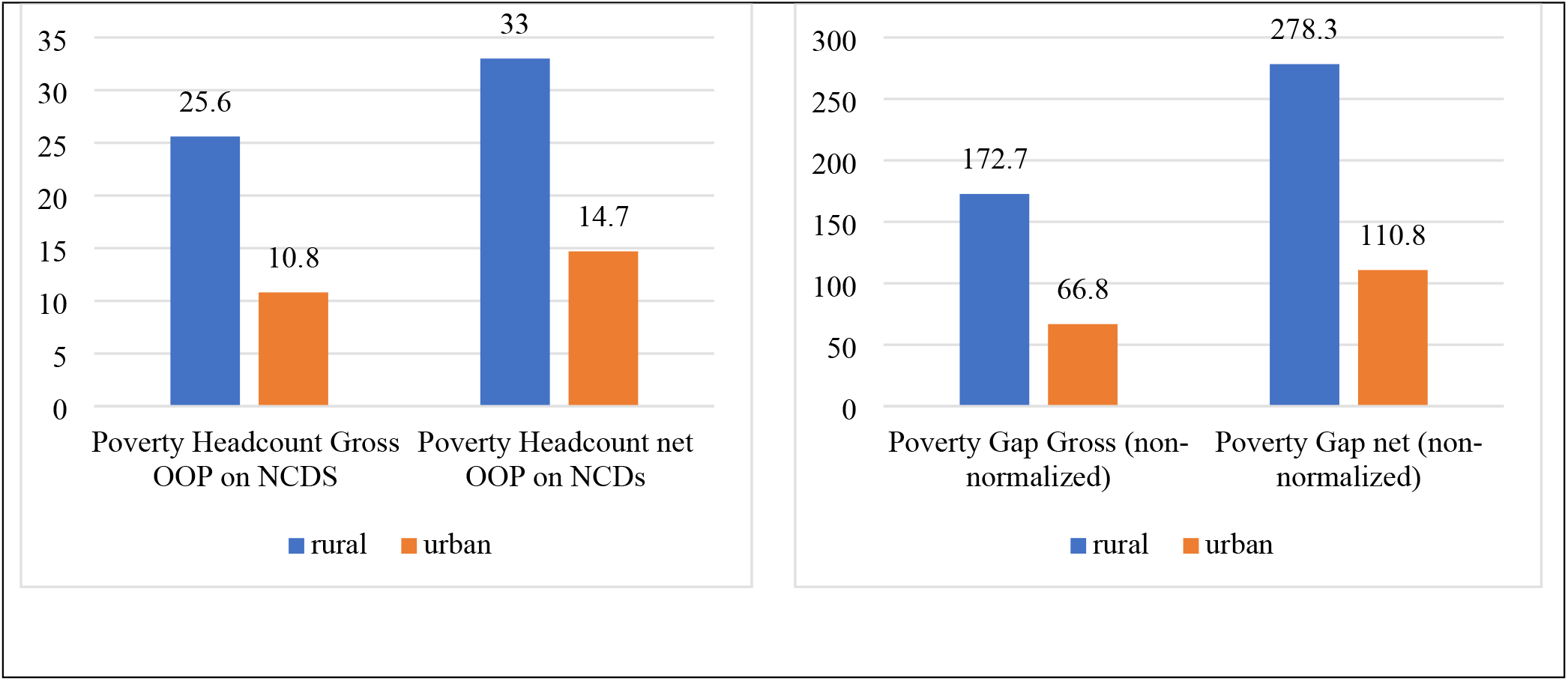
Impoverishment effect of OOP on NCDs by region.

### Socio-demographic and economic profile of households of the study

Table 2 presents descriptive statistics of the variables. Over three-fourths (83%) of the household heads were employed during 2018-19. Only 9 percent were female-headed families, and half of the heads were middle-aged (45 years). Most household heads (43%) had no education while only 7% had graduated. The age composition of the family showed that 80% of families had both children (aged <18 years) and older (>60 years) members implying a high dependency ratio within households, one-third of families had only older members present, and 14% had only children. More than three-fourths of households (84%) sought treatment for non-communicable diseases from public healthcare units. It may be due to the unaffordability of private healthcare services, which are mostly expensive and located in urban centers (Blum et al. 2018).

**Table 2.**
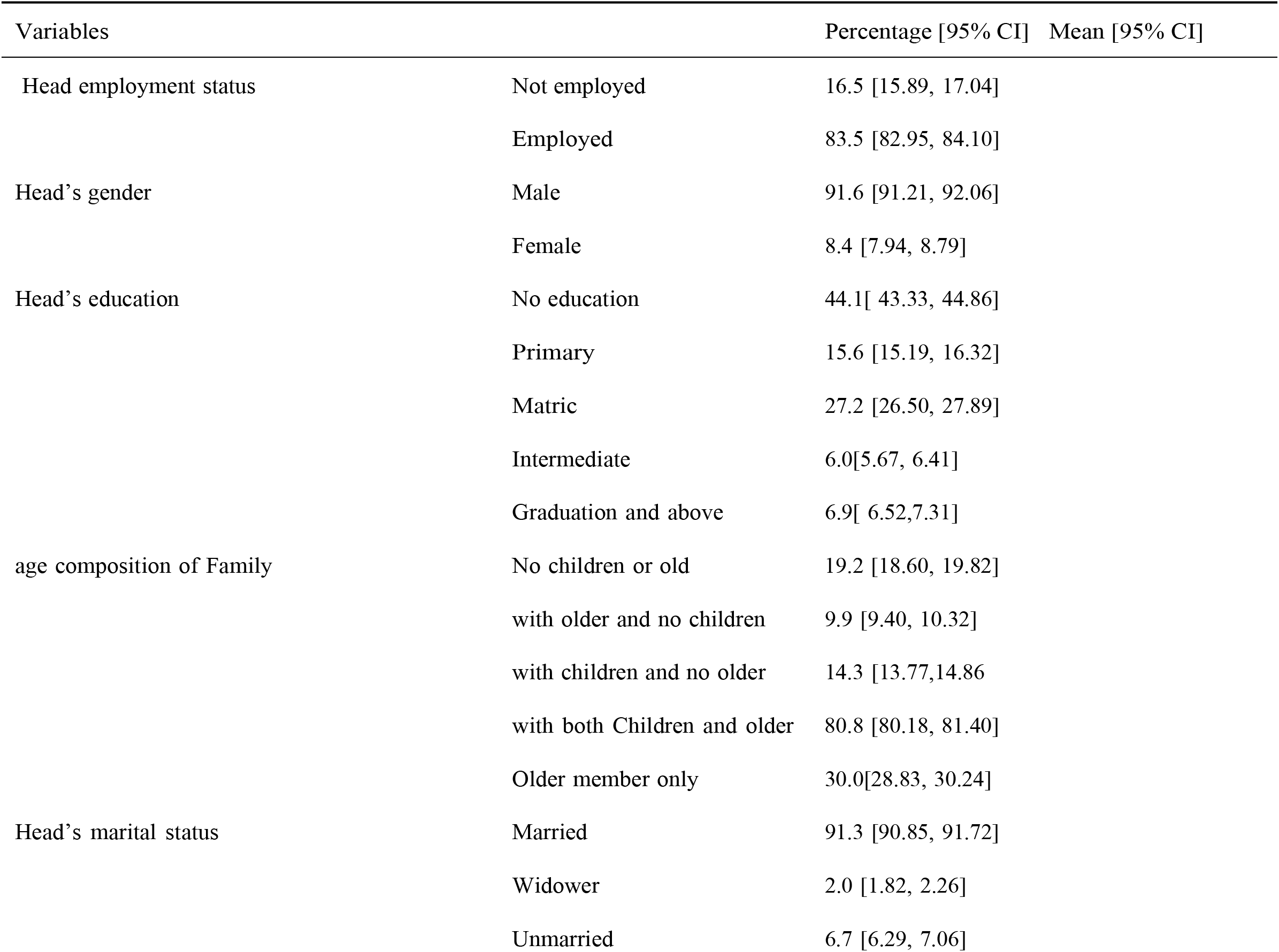
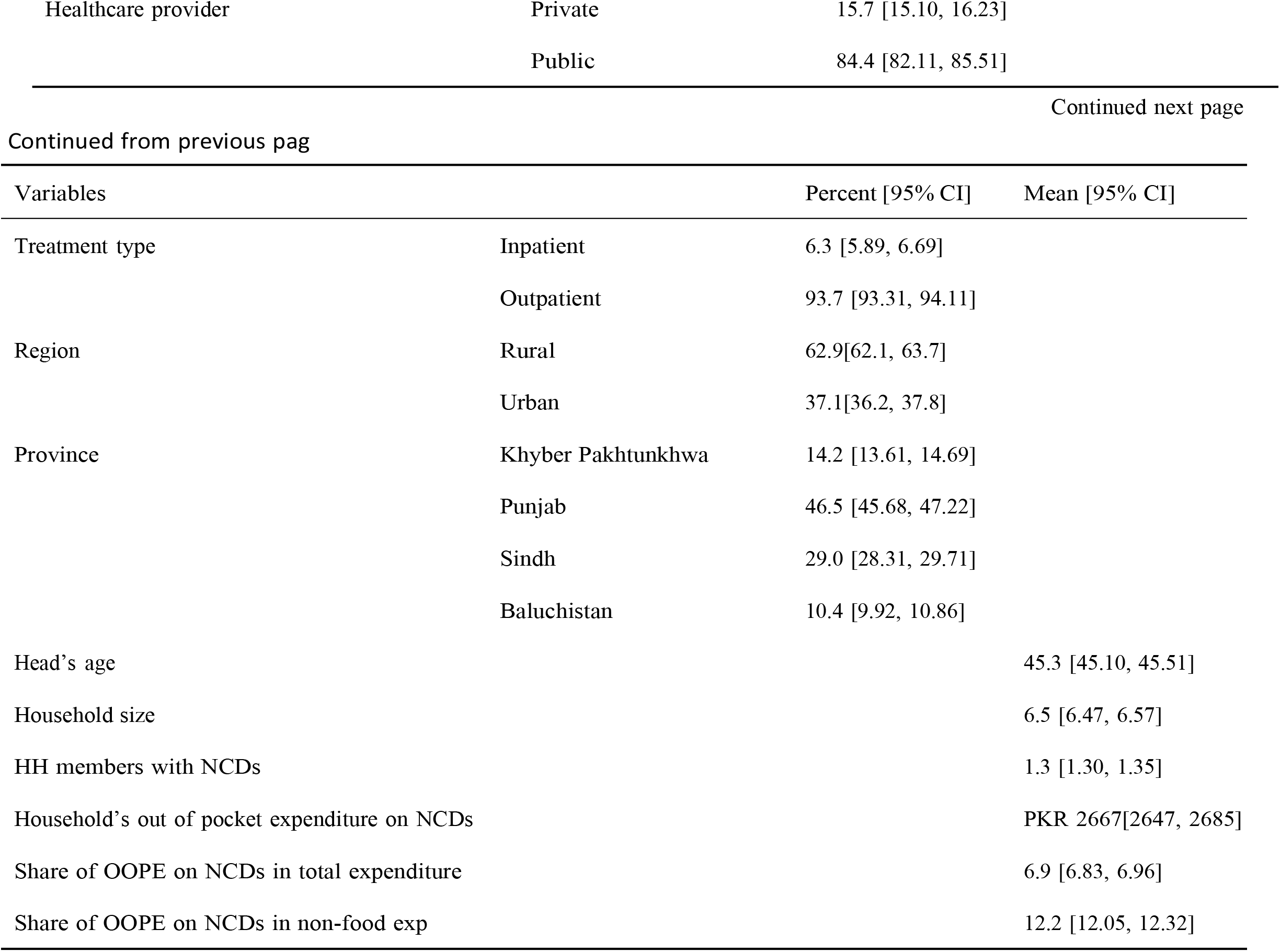
Descriptive statistics

In Pakistan, private healthcare providers are unregulated, resulting in costly treatments for cancer, cardiovascular disease, HIV, and others. As a result, middle- and lower income households are forced to seek treatment from public hospitals, which lack modern equipment, medicine, and staff (Hasan et al. 2022; Khan, AM., Ahmed, and Evans 2017; Wasay et al. 2014). Likewise, a higher proportion (90%) of households sought outpatient treatment (OPT), which reflects the need for healthcare financing covering OPT, particularly for the needy and poor. The average household’s OOP expenditure on NCDs was PKR 2,633 (USD 19) per month, and shares of OOP were 7% and 12% in the overall and non-food expenditure, respectively.

### Determinants of catastrophic health expenditure associated with non-communicable diseases

Table 3 shows estimates of the determinants of CHE. The family size had a positive and significant impact on CHE, and the magnitude of the effect was notably higher at the 15% and 20% thresholds. Moreover, family members with NCDs and the likelihood of CHE were positively related, and results were significant at all thresholds, ranging from 5% to 20%, see Table 3. Compared to the married head, the probability of CHE was lower among unmarried and widow/widower at all thresholds, implying a higher economic burden on families headed by married heads. The CHE on NCDs by gender showed that female-headed households were less likely to experience CHE on NCDs compared to male-headed.

**Table 3.**
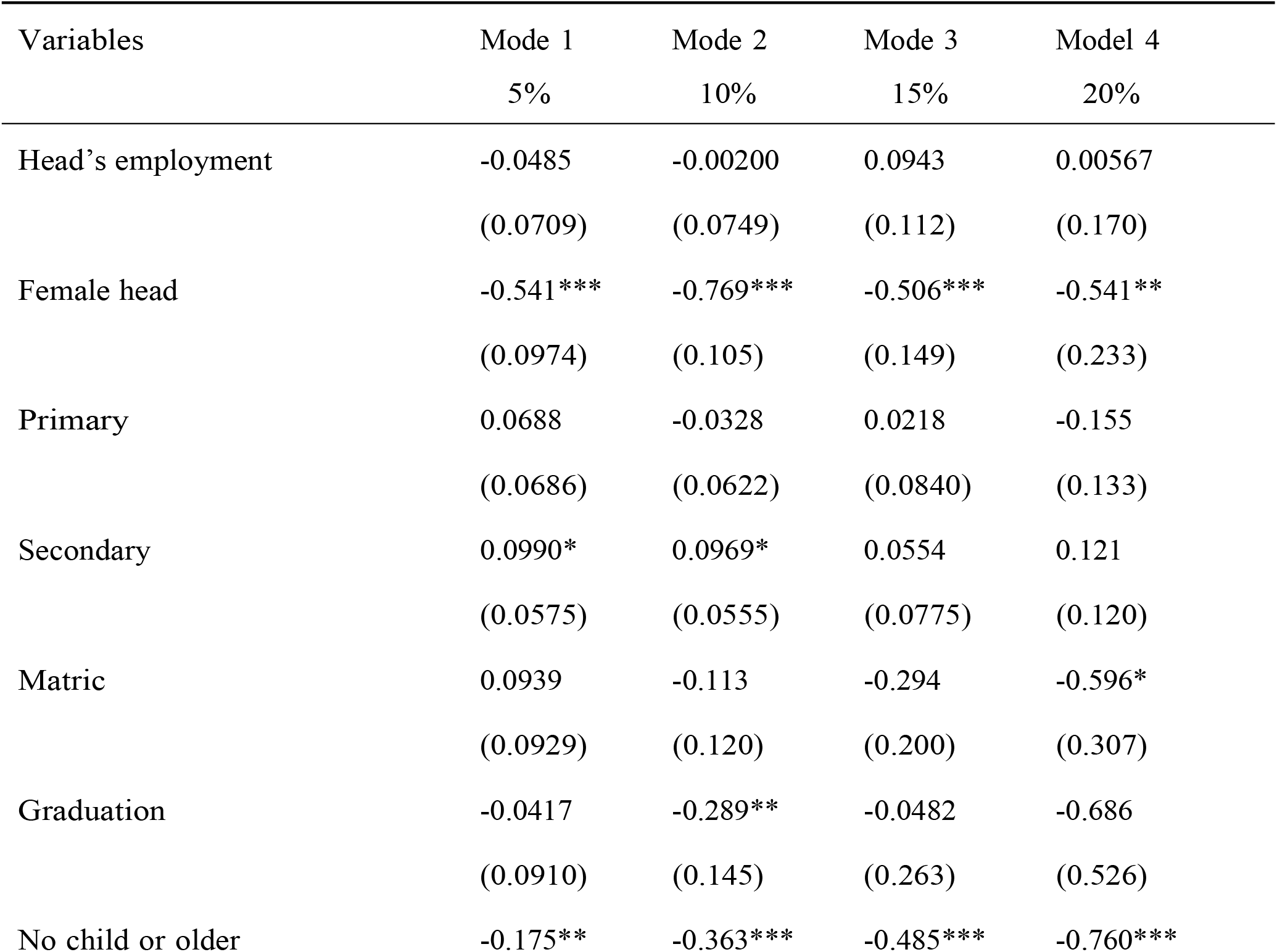
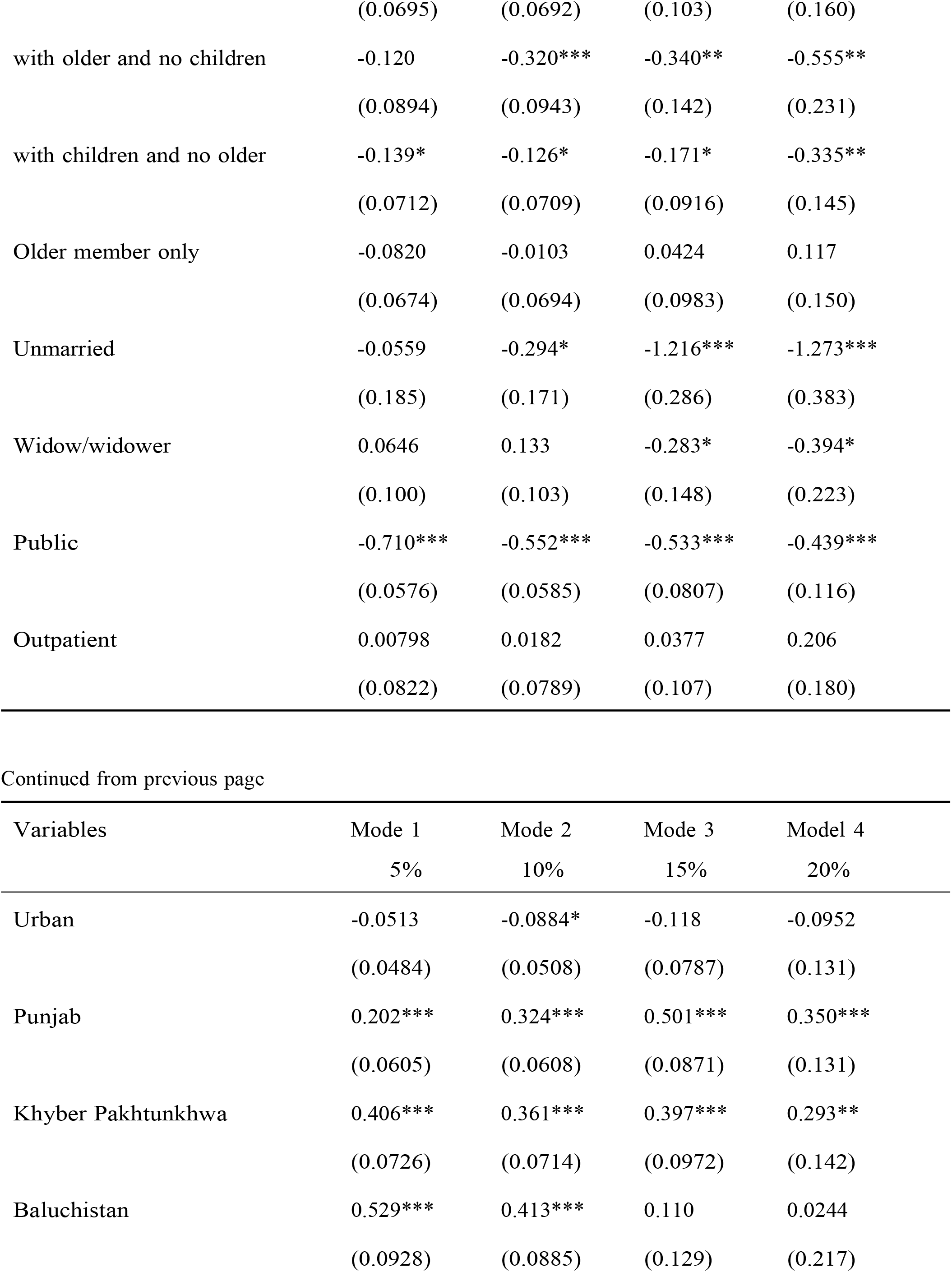
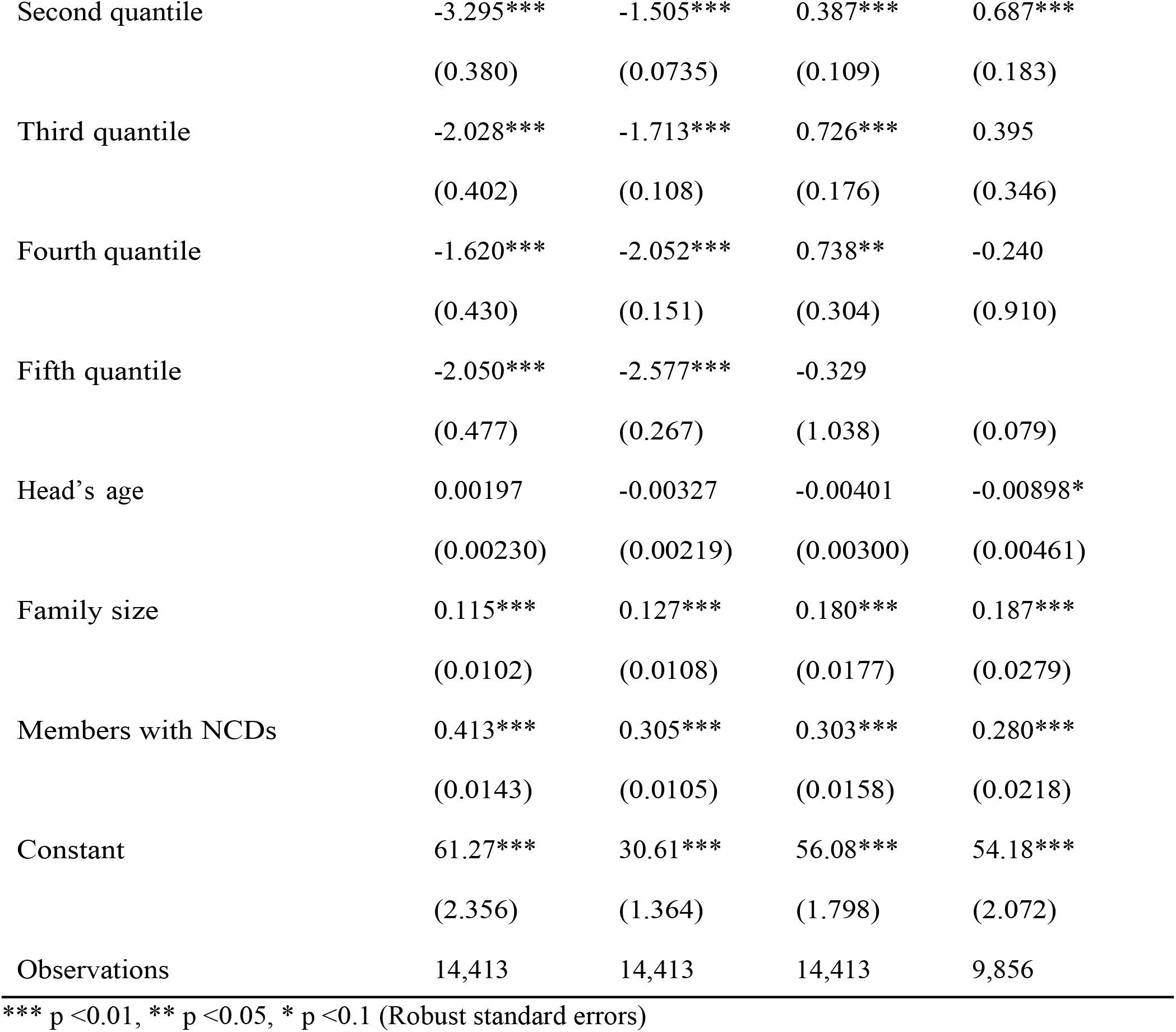
Determinants of catastrophic expenditure associated on non-communicable disease in Pakistan

The results were significant at all thresholds. In Pakistan, female-headed households are less common due to the patriarchal society. There are a few exceptions, where male members are not present, females take charge of the family. However, the pervasive gender inequality, cultural restrictions on women’s mobility, and low education among females are likely to contribute to the lower healthcare spending among female-headed households. Families headed by highly qualified persons (graduation and above) had a lower likelihood of CHE compared to families headed by persons with no education at all at a 10% threshold. However, the results were not significant for other thresholds.

The age composition of the household members indicates a higher probability of experiencing catastrophic health costs among families with children (less than 18 years of age) and older (above 60 years of age) adults at various thresholds. For example, compared to families with children and older members present, households with older members and no children had 28% lesser relative risk (exp^-0.32=0.72) of experiencing CHE at a 10% threshold, families with children but without older members were associated with 11.3%lower relative risk (exp^- 0.12=0.88). In contrast, families without both children and older members had 30% (exp^- 0.36=0.69) lower risk.

Compared to the private healthcare provider, families seeking healthcare from the public were less likely to encounter CHE on NCDs at all thresholds. The effect of household income (transformed into the log) on CHE is significant and negative at all thresholds. A positive difference in income (equivalent to increase in income) decreases the odds of CHE on non-communicable diseases. It implies that the economic burden of NCDs tends to be higher at a lower income.

Similarly, the likelihood of CHE was lower among higher-income quintiles compared to the lowest quintile, implying a higher incidence of CHE among lower-income groups. Moreover, a region-wise analysis indicates a lower likelihood of CHE in urban areas than the rural. It may be due to increased access to multiple healthcare providers and a lower incidence of NCDs. Across provinces, Punjab, KPK, and Baluchistan had a higher probability of CHE on NCDs than Sindh (reference category). The results were significant for all thresholds, except at 15% and 20% for Baluchistan.

### Impoverishing effects of catastrophic health expenditure

In this study, we calculated the incidence and intensity of poverty among households resulting from OOP spending on treating NCDs. The difference between the incidence of poverty before and after healthcare payments was used to determine the impact of OOP health expenditure on poverty. The poverty gap was calculated to quantify the degree to which OOP expenses caused the household to fall below the poverty line. Besides, normalized, and non-normalized poverty gaps were estimated to determine the cost needed to lift the individual out of poverty.

Table 4 shows results on the impoverishment effects of OOP NCDs expenditure. The poverty headcount was 20.5% without adjustments in OOP health expenditure and OOP expenditure on NCDs; the poverty headcount increased to 25% when OOP expenditure on healthcare were netted out of adult equivalent household expenditure and reached 27% after accounting for OOP spending on NCDs. The number of poor increased by 8 million (from 42.4 million to 51.3 million) due to OOP expenditure on health and by 13 million (from 42.4 million to 55.6) on account of OOP expenditure on NCDs in 2018-19, indicating gigantic impoverishment effect of OOP expenditure on NCDs in Pakistan. Correspondingly, the poverty gap attributable to OOP expenditure on health increased by 48%, amounting to PKR 196.3, expressed as a percentage of the poverty line. The poverty gap increased to 65% or PKR 219.1 (non-normalized) when health payments on NCDs were netted out of adult equivalent household expenditure. A region-wise pre-post analysis of the incidence and intensity of poverty indicates rural households experienced a higher burden of impoverishment compared to the urban when OOP expenditure on NCDs was netted out of adult household expenditure, see Figure 3.

**Table 4.**
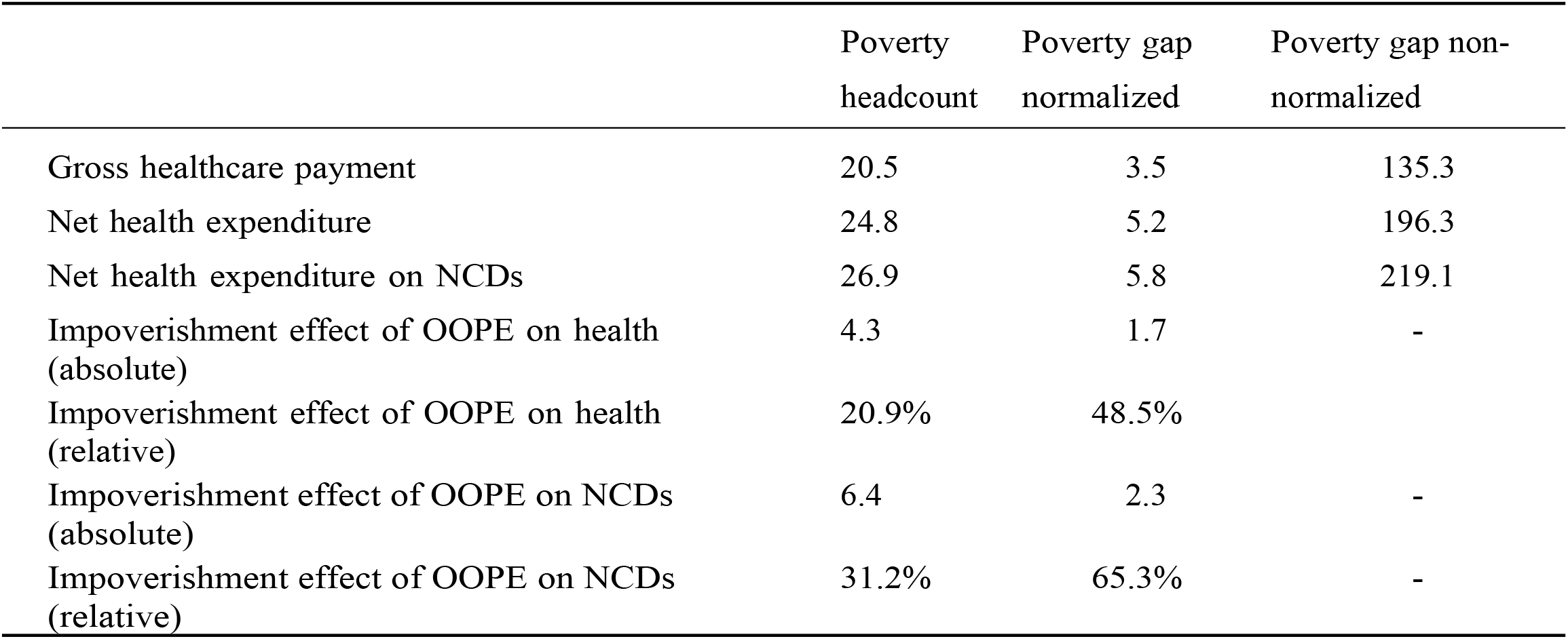
The impoverishment effect of OOPE on healthcare and non-communicable diseases in Pakistan in 2018-19

### Incidence and Intensity of catastrophic health expenditure due to non-communicable diseases

Table 5 provides estimates of the incidence and intensity of CHE on NCDs in 2018-19. The percentage of households experiencing CHE on NCDs falls from 60% to 3.5% as the threshold increases from 10% to 40%, implying fewer households encounter CHE at higher thresholds. Similarly, the overshoot increased from 10.4% to 12.3% when the threshold increased from 10% to 40%. However, the mean overshoot (MPO) increased to 48% when the threshold increased from 10% to 20% and dropped to 23% at the 40% threshold. Households who spent more than 10% of non-food expenditure spent 17.2% on average on getting NCD treatment. In comparison, those whose spending exceeded the 20% threshold of non-food expenditure had average spending of around 48% on NCDs.

**Table 5.**
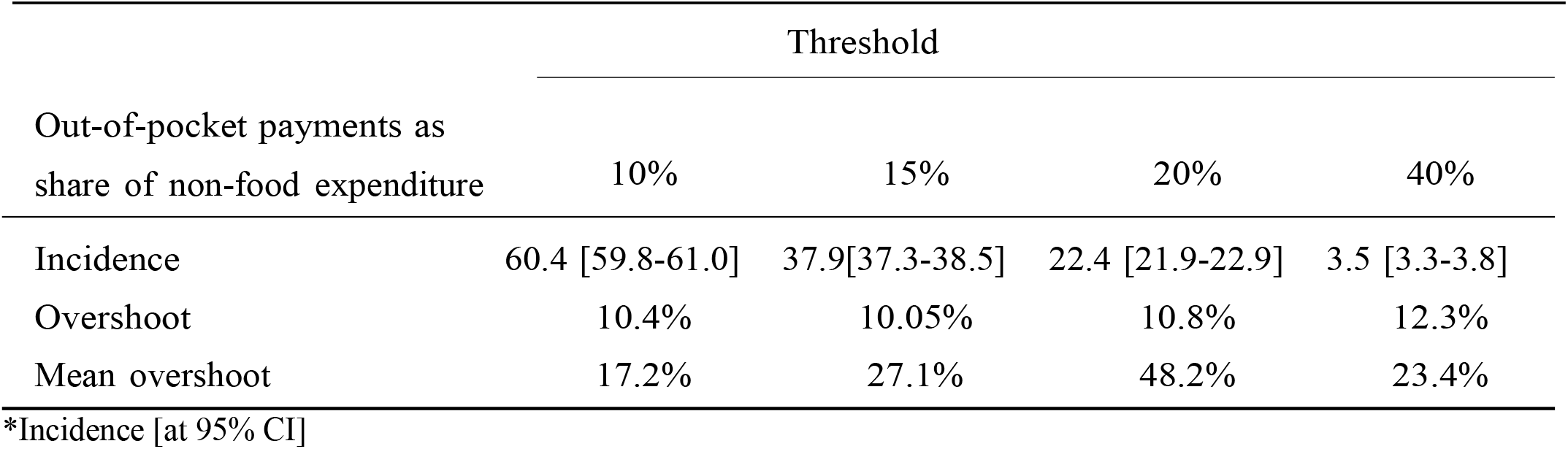
Incidence and intensity of catastrophic health expenditure associated with NCDs from nonfood expenditure in Pakistan

| Out-of-pocket payments as share of non-food expenditure | Threshold |  |  |  |
| --- | --- | --- | --- | --- |
|  | 10% | 15% | 20% | 40% |
| Incidence | 60.4 [59.8-61.0] | 37.9[37.3-38.5] | 22.4 [21.9-22.9] | 3.5 [3.3-3.8] |
| Overshoot | 10.4% | 10.05% | 10.8% | 12.3% |
| Mean overshoot | 17.2% | 27.1% | 48.2% | 23.4% |

\*Incidence [at 95% CI]

## Discussion

This study observed a higher incidence of OOP expenditure on NCDs in rural and in the lowest income quintile. In addition, OOP expenditure on NCDs were higher than those on healthcare for all income quintiles, reflecting the urgency to address the healthcare needs of people experiencing NCDs. Large families, members with NCDs and families with children and older members, male heads, heads with partners, and families using private healthcare treatment were more likely to incur catastrophic expenditure on NCDs.

Moreover, income and the likelihood of CHE were inversely related; higher income reduced the probability of CHE on NCDs and vice versa. Similar patterns were observed across income quintiles, as households at lower quintiles were more likely to spend higher on NCDs than those at higher quintiles. The impoverishment effect of OOP expenditure on NCDs indicates that 13 million more people (from 42.4 million to 55.8 million) fell into poverty in 2018-19 when OOP expenditure on NCDs were netted out from adult equivalent household expenditure. Similarly, the poverty gap increased from 3.5 to 5.6 (from PKR 135 to PKR 219.1) when we adjusted adult equivalent household expenditure for OOP expenditure on NCDs. Urban households experience a less impoverishment effect of CHE on NCDs than the rural.

Moreover, the gross poverty headcount and poverty gaps were higher among rural households. Rural households are generally resource-constrained in Pakistan and therefore rely heavily on the public healthcare system, which is not sufficient to accommodate the medical needs of the increasing rural population. For example, about 100,000 female health workers offer basic maternal and preventative childcare services, immunizations, and other services to about two-thirds people of the country (Islam, Malik, and Basaria 2002). However, the staff lacks professional training to provide care to patients with NCDs (Jafar et al. 2013). As a result, patients and their caretakers travel to urban centers to seek medical treatment by spending a hefty amount of money on transportation and rented lodgings.

The OOP health expenditure on NCDs can perpetuate poverty. A study in India observed an increase in impoverishment effect due to OOP expenditure on NCDs; 10% more people in 2014 and 12% in 2017-18 fell into poverty due to OOP expenses on NCDs. Further, 25% and 20% of those who were already poor fell deeper into poverty in 2014 and 2015, respectively (Verma, R. et al. 2021). Similarly, a study in Bangladesh established that poverty increased by 3.4% (from 37.8% to 41.1%) when OOP expenditure on healthcare were netted out from total household expenditure (Khan, AM. et al. 2017).

Moreover, the impoverishment effect deepens for the already-poor individuals. In Nepal, a longitudinal investigation into the Kala-azar affected households showed that 20–26% of non-poor people fell into poverty due to OOP expenditure on inpatient Kala-azar care. The poverty gap was 142% in the first year, which jumped to 184% in the following year for non-payment of loans to get medical treatment for Kala-azar. The squared poverty gap showed a similar trend, increasing from 560% in the first year to 3094% in the fifth year (Adhikari R., Maskay M., and Sharma P. 2009).

The impoverishing health expenditure have several adverse implications, such as reduced adherence to long-term therapy, abandonment of treatment, and impaired quality of life (Peters et al. 2008). Furthermore, unlike wealthy households, the poor are constrained in the choice of coping strategies, as they have to resort to using limited savings, selling assets, borrowing, and involving their children in the labor market, to tackle with NCDs related impoverishment (Margaret E Kruk, Goldmann, and Galea 2009). This connection, whereby a household’s spending on health causes that household to become even more impoverished, can result in a vicious cycle of ill health, lower productivity in the labor market, and poverty. Using 2010 as a base scenario, a study in Pakistan found cumulative production losses attributable to NCDs standing around USD 3·47 billion. The economic burden was projected to rise sharply from USD 152 million in 2010 to USD 296 million in 2025(Jafar et al. 2013).

Different denominators have been used in the literature to capture CHE, including nonfood expenditure, total expenditure, and income. In our study, the incidence of CHE on NCDs decreases as a measure of nonfood expenditure as we increase the threshold from 10% to 40%. Conversely, the intensity of the catastrophic or overshoot increased for every increase in the threshold ranging from 10%-40%. Households spent more than 10.8% beyond the 20% threshold of nonfood expenditure.

The mean overshoot among those spending beyond the 20% threshold of nonfood expenditure was 48.2%. Thus, these households spent 68.2%, a sum of overshoot and mean overshoot, of their non-food expenditure on the treatment of NCDs. Similar evidence from LMICs showed households were forced to pay as much as eight days’ worth of wages to purchase one month’s supply of only one of the multiple medicines required for the optimal treatment of cardiovascular disease (CVD) or diabetes (Cameron et al. 2009).

Our study showed a positive relationship between the probability of incurring catastrophic expenditure and households with NCDs afflicted members. The results confirm earlier evidence on the risk factors of CHE for selected 18 countries. After accounting for covariates, the study showed that the absolute risk of CHE was higher among households having members with NCDs than families without NCDs. However, the risk difference was the highest in low-middle-income countries (risk difference= 1.71%), followed by upper-middle-income (risk difference= 1.27%) and China (risk difference=7.52%). A similar pattern was observed in the impoverishment risk for upper-middle-income countries and China; UMICs (risk difference: 0.34%) and China (risk difference: 1.15%) (Murphy et al. 2020).

Similarly, a study in Bangladesh found that households with at least one member experiencing NCDs had more significant share of healthcare expenditure in the total expenditure than families without any such member. Moreover, households with NCDs had a 6.5% higher probability of incurring catastrophic expenditure, and an 85% increased likelihood of selling assets or borrowing from the informal market to meet with medical expenses than households without NCDs (Datta, K. et al. 2018).

In low-income-countries, including Pakistan, policies have been geared to integrate the programs aiming at improving the effectiveness of healthcare systems with economic growth policies, poverty alleviation plans, and social mobility and employment generation (Jafar et al. 2013; Jan et al. 2018). Despite the holistic approach, the increasing economic burden of NCDs in LMICs calls for extending the outreach of the financial against healthcare risks. There is a need to develop drastic measures to eliminate financial obstacles to uptake of preventive, curative, and support care services among NCD patients (Cameron et al. 2009; Jan et al. 2018; Kankeu et al. 2013). A healthcare system promoting adherence to cost-effective interventions designed to help the poor is crucial for achieving the sustainable development goal of poverty reduction (Jan et al. 2018).

## Data Availability

The study is available publicly from the Pakistan Household Integrated Economic Survey or Pakistan Social and Living Standards Measurement Survey. The data is available at reasonable request from the authors

## List of abbreviations

OOP: Out of pocket
OOPE: Out-of-pocket expenditure
NCD: Non-communicable disease
UHC: Universal health coverage
CHE: Catastrophic health expenditure
UMIC: Upper middle-income countries
LMIC: Lower middle-income countries
OPT: Outpatient treatment
SDG: Sustainable development goals
WHO: World Health Organization
HIES: Household integrated and economic health survey

